# Risk-Adapted Atrial Fibrillation Monitoring after Embolic Stroke of Undetermined Source: A Population-Based Study

**DOI:** 10.64898/2026.08.27.26361578

**Authors:** Julian Elbischger, Andreas Krainer, Tanja Ruprechter, Melanie Haidegger, Natalie Berger, Isra Hatab, Simon Fandler-Höfler, Martin Heine, Jan Jagiello, Herbert Koller, Stefan Lilek, Sai Pavan Kumar Veeranki, Christian Enzinger, Martin Manninger, Egbert Hubertus Bisping, Daniel Scherr, Thomas Gattringer, Markus Kneihsl

**Author notes:** Dr. Elbischger and Dr. Krainer contributed equally to the manuscript. **Corresponding author:** Markus Kneihsl, MD, PhD Department of Neurology, Medical University of Graz Auenbruggerplatz 22, A-8036 Graz, Austria Phone: +43/316/385/82984 Mail.

## Abstract

**Background:** Atrial fibrillation detected after stroke (AFDAS) is frequently diagnosed after embolic stroke of undetermined source (ESUS) and has important implications for secondary stroke prevention. Although prediction scores have been proposed to identify patients at increased risk of AFDAS, prospective evidence supporting their implementation to guide rhythm monitoring in routine clinical practice is limited.

**Methods:** In this prospective, population-based implementation cohort study, adults with ESUS were enrolled between January 2022 and December 2024 across all stroke centers in Styria, Austria. The Graz AF Risk Score was prospectively implemented as part of a risk-adapted diagnostic pathway for cardiac rhythm monitoring. Patients with a score ≥4 were recommended for implantable loop recorder monitoring, whereas monitoring in those with scores <4 remained at the treating physician’s discretion. The primary outcome was AFDAS detection; recurrent ischemic stroke and recurrent stroke etiology were secondary outcomes.

**Results:** Among 784 patients (median age 73 years [IQR 64–80], 45.7% women), AFDAS was detected in 166 patients (21.2%) during a median follow-up of 26.3 months (IQR 20–34). AFDAS detection was substantially higher in patients with a Graz AF Risk Score ≥4 than <4 (38.1% vs. 3.9%; p<0.001). After adjustment for age, sex and ILR monitoring, a score ≥4 independently predicted AFDAS (HR 6.3, 95% CI 3.5–11.2; p<0.001) and recurrent ischemic stroke (HR 2.2, 95% CI 1.1–4.1; p=0.023). Only one recurrent stroke in patients with a score <4 was attributable to atrial fibrillation (AF) (1/18, 5.6%).

**Conclusions:** Prospective implementation of the Graz AF Risk Score identified patients with ESUS at markedly different risks of AFDAS. A Graz AF Risk Score ≥4 was also independently associated with recurrent ischemic stroke. These findings support a risk-adapted approach to cardiac rhythm monitoring after ESUS.

## Introduction

Embolic stroke of undetermined source (ESUS) represents a major challenge in secondary stroke prevention, as the underlying mechanism remains unresolved despite standardized diagnostic evaluation. ^1–3^ Occult atrial fibrillation (AF) is a clinically actionable underlying mechanism in a substantial subset of ESUS patients, with AF first detected after stroke (AFDAS) in 16–30% during follow-up. ^4–6^

Notably, empirical anticoagulation strategies in ESUS have consistently failed, with randomized trials showing no benefit of direct oral anticoagulation over antiplatelet therapy ^7–10^, even among patients with evidence of atrial cardiomyopathy. ^10^

These findings underscore the biological heterogeneity of ESUS and suggest that precise identification of occult AF, rather than empiric treatment escalation, may be central to improving secondary prevention. Accordingly, prolonged cardiac rhythm monitoring has become a key diagnostic strategy after ESUS. ^11^

However, broad implementation of intensive monitoring, particularly with implantable loop recorders (ILRs), remains constrained by cost and logistics and it remains unclear whether detection of low-burden AF translates into improved clinical outcomes. ^12^ The clinical challenge is therefore to identify those patients most likely to benefit from resource-intensive monitoring while avoiding low-yield investigation in others. In this context, several clinical prediction models have been proposed to improve AFDAS detection ^13–23^, but their role in guiding monitoring intensity remains uncertain, and robust evidence supporting pragmatic risk-guided implementation in routine clinical care is lacking. ^13–28^

In this prospective, population-based study across all stroke centers in the federal state of Styria, Austria, we evaluated the Graz AF Risk Score within a real-world, risk-adapted diagnostic pathway for predicting AFDAS in ESUS patients. We further examined whether the score was associated with stroke recurrence and recurrent stroke mechanisms during long-term follow-up.

## Methods

### Study design and patient population

The Graz AF Risk Score, previously developed to predict AFDAS following cryptogenic stroke ^23^, was prospectively implemented within a structured diagnostic pathway across all five neurology departments providing acute stroke care in the federal state of Styria, Austria, which serve a population of 1.2 million inhabitants.

All consecutive patients with acute ischemic stroke between 2022 and 2024 were eligible if classified as ESUS after a thorough diagnostic workup, which included neuroimaging (computed tomography and/or magnetic resonance imaging), neurosonography of extra- and intracranial arteries, transthoracic and/or transesophageal echocardiography, and at least 72 hours of ECG-monitoring according to current guidelines. ^29^ ESUS was defined according to established criteria, including the absence of (1) ≥50% stenosis in arteries supplying the infarct area, (2) major-risk cardioembolic sources, and (3) other specific causes despite a standardized diagnostic evaluation. ^29^ The study is reported in accordance with the Strengthening the Reporting of Observational Studies in Epidemiology (STROBE) guidelines. ^30^

### Diagnostic pathway

The Graz AF Risk Score was prospectively calculated in all eligible patients with ESUS according to the originally published criteria as part of routine care (for details see **Supplemental Figure 1**). ^23^ Informed by the derivation study, Graz AF Risk Scores ≥4 were considered suggestive of increased AF risk and were recommended for intensified cardiac rhythm monitoring via ILR. ^23^ However, monitoring decisions remained at the discretion of the treating physician, and patients with lower scores <4 could also receive an ILR. In patients in either risk-score category who did not undergo ILR implantation, standard rhythm surveillance included serial pulse assessments and repeated ECGs during hospitalization, as well as ambulatory Holter monitoring for up to 7 days. Patients with confirmed AFDAS received oral anticoagulation according to current guidelines. ^31^ The diagnostic pathway is illustrated in **Figure 1**.

**Figure 1.**
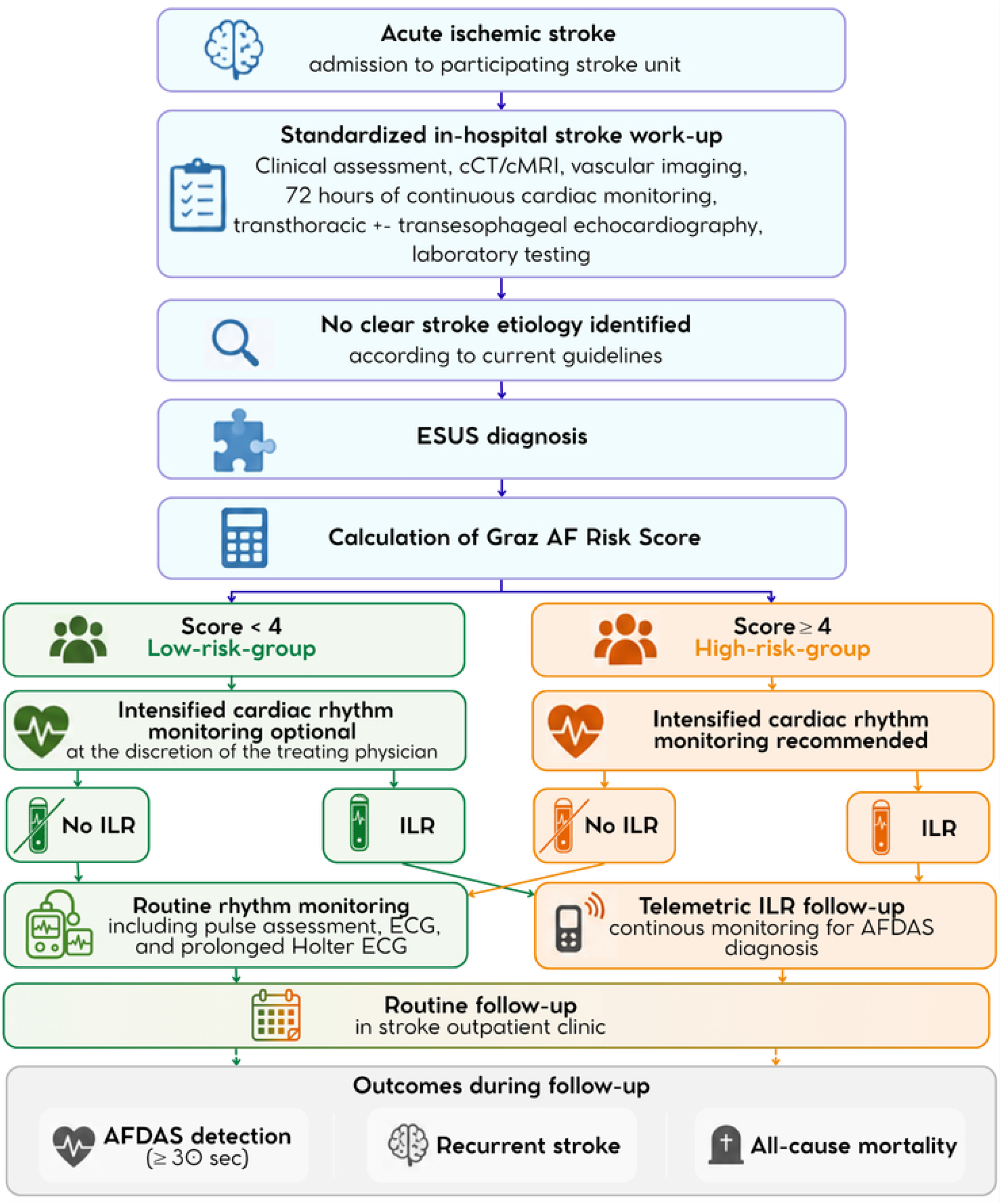
Structured diagnostic pathway and implementation of the Graz AF Risk Score in patients with ESUS Patients were prospectively stratified according to their Graz AF Risk Score. Scores ≥4 were considered suggestive of increased AF risk and prompted intensified rhythm monitoring, including 7-day Holter ECG and/or implantable loop recorder (ILR) implantation at the discretion of the treating physician. Patients with lower scores could also undergo prolonged rhythm monitoring. Abbreviations: AF, atrial fibrillation; AFDAS, atrial fibrillation detected after stroke; cCT, cranial computed tomography; cMRI, cranial magnetic resonance imaging; ECG, electrocardiogram; ESUS, embolic stroke of undetermined source; ILR, implantable loop recorder.

### Data Sources and Collection

Patient data were prospectively captured through the Austrian Stroke Unit Registry. ^32^ In addition, a dedicated electronic case documentation framework was implemented for all patients with ESUS to ensure systematic collection of study-specific variables relevant to AF risk stratification, including echocardiographic, electrocardiographic, N-terminal pro–B-type natriuretic peptide (NT-proBNP), and monitoring-related data (**Supplemental Figure 1**).

### Outcomes and Follow-up

The primary outcome of this study was the diagnosis of AFDAS, defined as ≥30 seconds of AF or atrial flutter, in accordance with current guideline recommendations, and documented by ECG, Holter monitoring, or ILR. ^31^ Secondary outcomes were recurrent ischemic stroke, recurrent ischemic stroke etiology and all-cause mortality.

Routine follow-up at 3 months was performed through in-person or telephone assessment. Long-term follow-up comprised the period from the index stroke until January 31, 2026, with administrative censoring at that date. Longitudinal outcome ascertainment was strengthened through integration of multiple complementary data sources, including the regional public healthcare network, which provides acute stroke care across the federal state of Styria, and the Austrian electronic health record system (ELGA), enabling capture of recurrent cerebrovascular events, clinically diagnosed AFDAS, and oral anticoagulation initiation across the study region.

### Statistical analysis

All data were analyzed using Statistical Package for the Social Sciences (SPSS - Version 29). Continuous variables are presented as median and interquartile range (IQR), and categorical variables as counts and percentages.

Baseline characteristics, score components, follow-up variables, and clinical outcomes were compared between patients with and without AFDAS, by ILR implantation status, and between predefined Graz AF Risk Score groups (high risk: ≥4 points; low risk: <4 points). Continuous variables were compared using the Mann–Whitney U test, while categorical variables were compared using the chi-square test or Fisher’s exact test. Associations between baseline characteristics and the continuous Graz AF Risk Score were assessed using Spearman’s rank correlation coefficient.

Time-to-event outcomes (AFDAS detection and recurrent ischemic stroke) were analyzed using Kaplan–Meier estimates and Cox proportional hazards regression. Time to event was defined as the interval from the index stroke to the first documented occurrence of the respective outcome.

To assess the independent association of the Graz AF Risk Score with AFDAS detection and recurrent ischemic stroke, multivariable Cox regression analyses were performed including age, sex, and ILR implantation as covariates.

The discriminative performance of the Graz AF Risk Score for AF prediction was evaluated using receiver operating characteristic (ROC) curve analysis. Sensitivity, specificity, positive predictive value, negative predictive value, and the Youden index were calculated for each score threshold.

A prespecified sensitivity analysis restricted to patients undergoing ILR implantation was performed analogously to the primary analysis. A two-sided p-value <0.05 was considered statistically significant for all analyses.

The study period (January 2022 through December 2024) and the administrative censoring date (January 31, 2026) were prespecified before data analysis. No formal sample size calculation was performed, as all consecutive patients with ESUS documented between 2022 and 2024 were included. The sample size was therefore determined by the available real-world cohort. There were minimal missing data for key study variables; analyses were therefore performed using a complete-case approach. A study flow diagram detailing patient inclusion and risk stratification is provided in **Figure 1**.

### Ethics

The study was conducted in accordance with the principles of the Declaration of Helsinki. Ethical approval was obtained from the Ethics Committee of the Medical University of Graz (REC number 29–285 ex 16/17).

## Results

Among 7336 patients with acute ischemic stroke during the study period, 784 (10.7%) fulfilled ESUS criteria and were included in the analysis (median age, 73 years [IQR, 64-80], 45.7% women). Median follow-up duration was 26.3 months (IQR, 20-34). During follow-up, AFDAS occurred in 166 patients (21.2%), with a median time to detection of 6 months (IQR, 1-12).

Compared with patients without AFDAS, those with AFDAS were older (>75 years: 65.7% vs 34.5%), more frequently had prior cortical or cerebellar infarction (35.5% vs 17.3%), ischemic stroke occurring despite ongoing antiplatelet therapy (39.8% vs 21.7%), left atrial enlargement (28.9% vs 13.3%), elevated NT-proBNP levels (>505pg/ml) (36.1% vs 18.4%), and increased supraventricular ectopy on 24-hour Holter monitoring (≥125 supraventricular premature beats: 63.9% vs 35.0%) (all p<0.001; **Table 1**).

**Table 1.** Baseline characteristics, components of the Graz AF Risk Score, and clinical outcomes in the overall cohort and stratified by atrial fibrillation status.

| Characteristics | Overall<br>(n=784) | AFDAS<br>(n=166) | No AFDAS<br>(n=618) | p* |
| --- | --- | --- | --- | --- |
| Baseline |  |  |  |  |
| Age, years (median, IQR) | 73 (64-80) | 79 (72-83) | 70 (62-79) | <.001 |
| Age>75 (n, %) | 322 (41.1) | 109 (65.7) | 213 (34.5) | <.001 |
| Age 60-75 (n, %) | 355 (45.3) | 51 (30.7) | 304 (49.2) | <.001 |
| Women (n, %) | 358 (45.7) | 73 (44.0) | 285 (46.1) | .623 |
| Old cortical / cerebellar<br>Infarct (n, %) | 166 (21.2) | 59 (35.5) | 107 (17.3) | <.001 |
| Multi- territory brain infarct (n, %) | 156 (19.9) | 36 (21.7) | 120 (19.4) | .516 |
| Infarct despite antiplatelet therapy (n, %) | 200 (25.5) | 66 (39.8) | 134 (21.7) | <.001 |
| EF <40% (n, %) | 15 (1.9) | 5 (3.0) | 10 (1.6) | .244 |
| EF 40%-50% (n, %) | 55 (7.0) | 20 (12.0) | 35 (5.7) | .004 |
| Atrial enlargement (n, %) | 130 (16.6) | 48 (28.9) | 82 (13.3) | <.001 |
| E/e`>12 (n, %) | 55 (7.0) | 18 (11.3) | 37 (6.0) | .030 |
| SPB on ECG at admission (n, %) | 43 (5.5) | 19 (11.4) | 24 (3.9) | <.001 |
| SPB>125 over 24h (n, %) | 322 (41.1) | 106 (63.9) | 216 (35.0) | <.001 |
| Atrial run ≥20 beats (n, %) | 44 (5.6) | 20 (12.0) | 24 (3.9) | <.001 |
| NT-proBNP>505 pg/ml (EF>50%)<br>(n, %) | 174 (22.2) | 60 (36.1) | 114 (18.4) | <.001 |
| NT-proBNP>505 pg/ml (EF<50%)<br>(n, %) | 36 (4.6) | 13 (7.8) | 23 (3.7) | .025 |
| Graz AF Risk Score (median, IQR) | 4 (2-5) | 6 (4-7) | 3 (2-5) | <.001 |
| Graz AF Risk Score ≥4 (n, %) | 396 (50.5) | 151 (91.0) | 245 (39.6) | <.001 |
| ILR (n, %) | 215 (27.4) | 102 (61.4) | 113 (18.3) | <.001 |
| Follow-up |  |  |  |  |
| Follow up, months (median, IQR) | 26.3 (20-34) | 30.1 (22-37) | 25.7 (19-33) | .001 |
| AFDAS (n, %) | 166 (21.2) | - | - |  |
| Time to AF, months (median, IQR) | 6 (1-12) | 6 (1-12) | - | - |
| Recurrent stroke (n, %) | 70 (8.9) | 23 (13.9) | 47 (7.6) | .012 |
| Myocardial infarction (n, %) | 13 (1.7) | 7 (4.2) | 6 (1.0) | .004 |
| GI bleeding (n, %) | 10 (1.3) | 4 (2.4) | 6 (1.0) | .142 |
| All cause mortality (n, %) | 31 (4.0) | 12 (7.2) | 19 (3.1) | .015 |

AFDAS, atrial fibrillation detected after stroke; ILR, implantable loop recorder; EF, ejection fraction; SPB, supraventricular premature beats; ECG, electrocardiogram; NT-proBNP, n-terminal pro-brain natriuretic peptide; IQR, interquartile range; AF, Atrial fibrillation; GI, gastrointestinal
\* Comparison between patients with and without AFDAS

### Graz AF Risk Score and AFDAS

Patients with AFDAS had higher Graz AF Risk Scores than those without (median, 6 [IQR, 4-7] vs 3 [IQR, 2-5]; p<0.001). At the predefined threshold of ≥4, AFDAS was identified in 151 of 396 high-risk patients (38.1%) compared with 15 of 388 low-risk patients (3.9%), corresponding to an absolute risk difference of 34.2 percentage points (p<0.001). Median time to AFDAS diagnosis was similar between groups (median, 6 months [IQR, 1-12] vs 6 [IQR, 2-11]; p=0.937; **Supplemental Table 1**). Kaplan–Meier analysis demonstrated early and sustained separation of cumulative incidence curves with a higher cumulative incidence of AFDAS in the high-risk group (log-rank p<0.001; **Figure 2**). After adjustment for age, sex, and ILR monitoring, a Graz AF Risk Score ≥4 remained independently associated with AFDAS (HR, 6.3; 95% CI 3.5-11.2, p<0.001; **Supplemental Table 2**).

**Figure 2.**
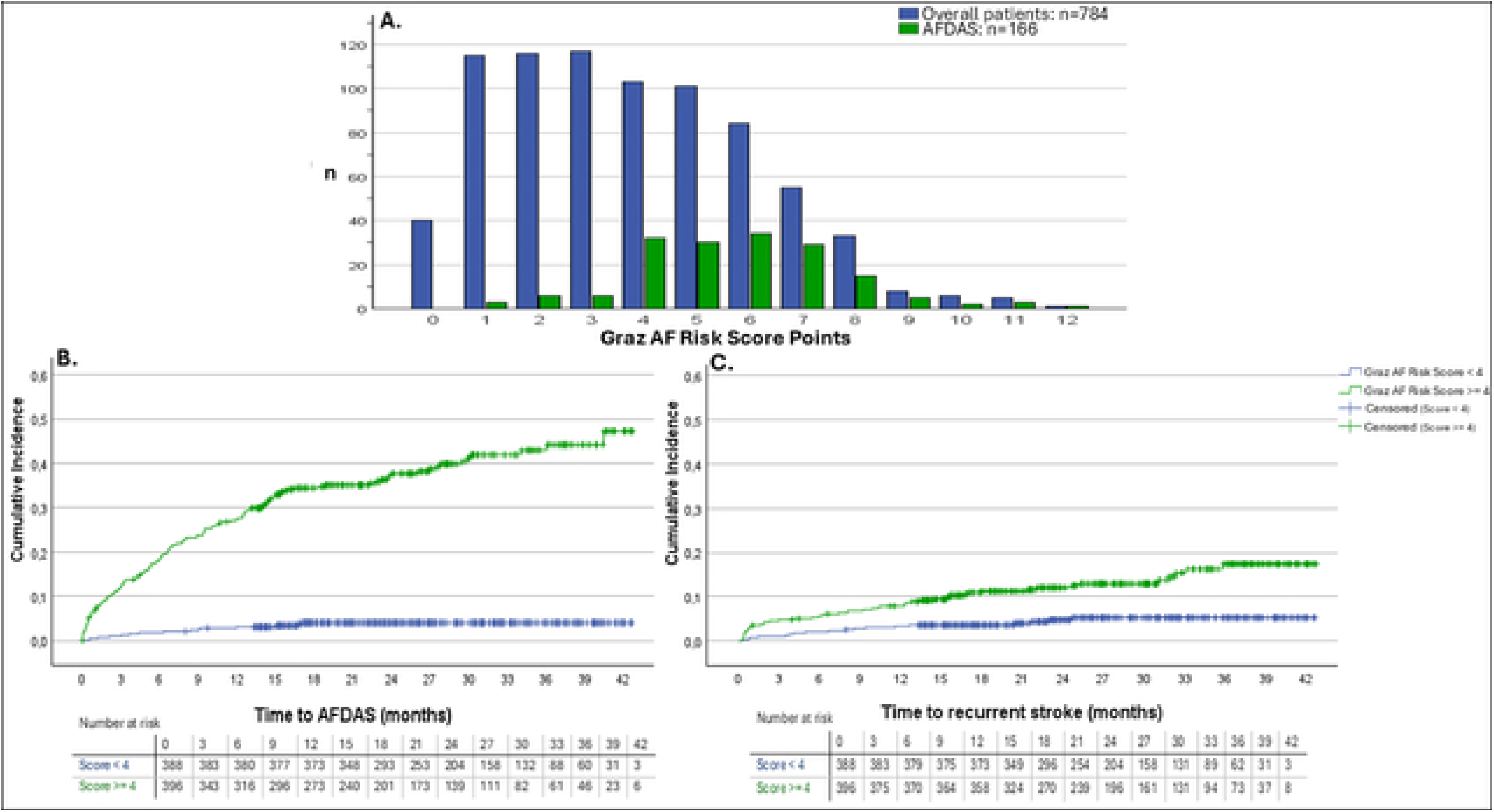
Graz AF Risk Score distribution and cumulative incidence of atrial fibrillation detected after stroke and recurrent ischemic stroke A, Distribution of Graz AF Risk Scores in the overall cohort and among patients with AFDAS. B, Cumulative incidence of AFDAS according to Graz AF Risk Score category (<4 vs ≥4). C, Cumulative incidence of recurrent ischemic stroke according to Graz AF Risk Score category (<4 vs ≥4). Numbers at risk are shown below the time-to-event plots. Groups were compared using the log-rank test. Abbreviations: AFDAS, atrial fibrillation detected after stroke; CI, confidence interval.

Discriminative performance of the Graz AF Risk Score for AFDAS was good, with an AUC of 0.79 (95% CI, 0.76–0.83) (**Table 2 and Supplemental Figure 3**). At the predefined threshold of ≥4, sensitivity was 91.0%, specificity was 60.4% and negative predictive value 96.1%. The number needed to intensively monitor to detect one case of AFDAS was 2.9. Detailed threshold-specific performance metrics are provided in **Table 2**.

**Table 2.**
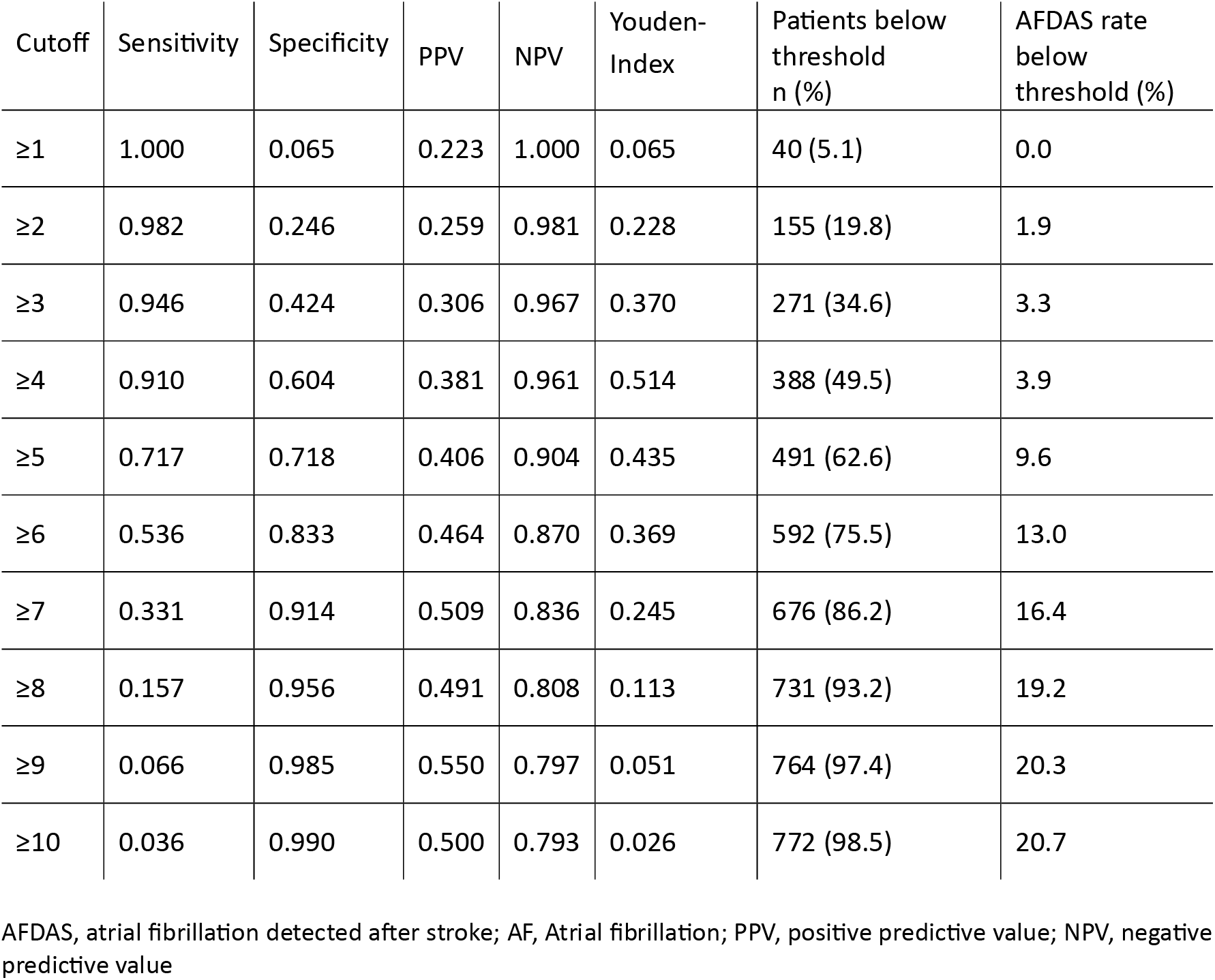
Diagnostic performance of the Graz AF Risk Score for atrial fibrillation detection across different cutoff values in the overall cohort.

| Cutoff | Sensitivity | Specificity | PPV | NPV | Youden-Index | Patients below threshold n (%) | AFDAS rate below threshold (%) |
| --- | --- | --- | --- | --- | --- | --- | --- |
| ≥1 | 1.000 | 0.065 | 0.223 | 1.000 | 0.065 | 40 (5.1) | 0.0 |
| ≥2 | 0.982 | 0.246 | 0.259 | 0.981 | 0.228 | 155 (19.8) | 1.9 |
| ≥3 | 0.946 | 0.424 | 0.306 | 0.967 | 0.370 | 271 (34.6) | 3.3 |
| ≥4 | 0.910 | 0.604 | 0.381 | 0.961 | 0.514 | 388 (49.5) | 3.9 |
| ≥5 | 0.717 | 0.718 | 0.406 | 0.904 | 0.435 | 491 (62.6) | 9.6 |
| ≥6 | 0.536 | 0.833 | 0.464 | 0.870 | 0.369 | 592 (75.5) | 13.0 |
| ≥7 | 0.331 | 0.914 | 0.509 | 0.836 | 0.245 | 676 (86.2) | 16.4 |
| ≥8 | 0.157 | 0.956 | 0.491 | 0.808 | 0.113 | 731 (93.2) | 19.2 |
| ≥9 | 0.066 | 0.985 | 0.550 | 0.797 | 0.051 | 764 (97.4) | 20.3 |
| ≥10 | 0.036 | 0.990 | 0.500 | 0.793 | 0.026 | 772 (98.5) | 20.7 |

AFDAS, atrial fibrillation detected after stroke; AF, Atrial fibrillation; PPV, positive predictive value; NPV, negative predictive value

### Sensitivity analysis

As intended within the risk-adapted diagnostic pathway, ILR-monitoring was preferentially focused on higher-risk patients: Overall, 215 patients (27.4%) underwent ILR implantation at a median of 22 days (IQR, 8-77) after the index event, with substantially higher implantation rates in the high-risk than in the low-risk group (189/396 [47.7%] vs 26/388 [6.7%]; p<0.001). (**Supplemental Table 1 and 3**) Reasons for not undergoing ILR implantation among high-risk patients are summarized in **Supplemental Table 4**. The most common reasons were patient refusal (33.2%) and a decision by the treating physician against ILR implantation (55.6%), most commonly because of severe post-stroke disability or multimorbidity.

Among patients undergoing ILR implantation, AFDAS occurred in 99 of 189 high-risk patients (52.4%) compared with 3 of 26 low-risk patients (11.5%) (p<0.001), yielding one AFDAS diagnosis for every 1.9 ILRs implanted in high-risk patients. After adjustment for age and sex, a Graz AF Risk Score ≥4 remained independently associated with earlier AFDAS detection (HR 5.4, 95% CI 1.7–17.1; p=0.004). (**Supplemental Table 3 and 5**) The distribution of AFDAS events across individual Graz AF Risk Score categories according to ILR implantation is shown in **Supplemental Figure 2**.

### Recurrent ischemic stroke

During follow-up, recurrent ischemic stroke occurred in 70 patients (8.9%), with a median time to recurrence of 7.5 months (IQR, 1-15). Recurrent stroke was more frequent among patients with AFDAS than among those without (13.9% vs 7.6%; p=0.012), and among patients classified as high risk compared with low risk by the Graz AF Risk Score (13.1% vs 4.6%; p=0.001). Kaplan–Meier analysis demonstrated early and sustained separation of recurrent stroke risk between high- and low-risk groups (log-rank p<0.001; **Figure 2)**. In multivariable Cox regression analysis adjusted for age, sex, and ILR implantation, a Graz AF Risk Score ≥4 remained independently associated with recurrent ischemic stroke (HR 2.2; 95% CI 1.1-4.1; p=0.023). (**Supplemental Table 6**)

Stroke mechanisms at recurrence remained heterogeneous. While cardioembolism was the most frequent stroke etiology in the Graz AF Risk Score based high-risk group (34.6%), macro- and microangiopathy were most prevalent causes in the low-risk group (33.3% each). Of note, among patients with a Graz AF Risk Score <4 points, only one individual was diagnosed with AF as the underlying etiology of the recurrent ischemic stroke (5.6%) (**Table 3**, **Figure 3 and Supplemental Table 7**).

**Figure 3.**
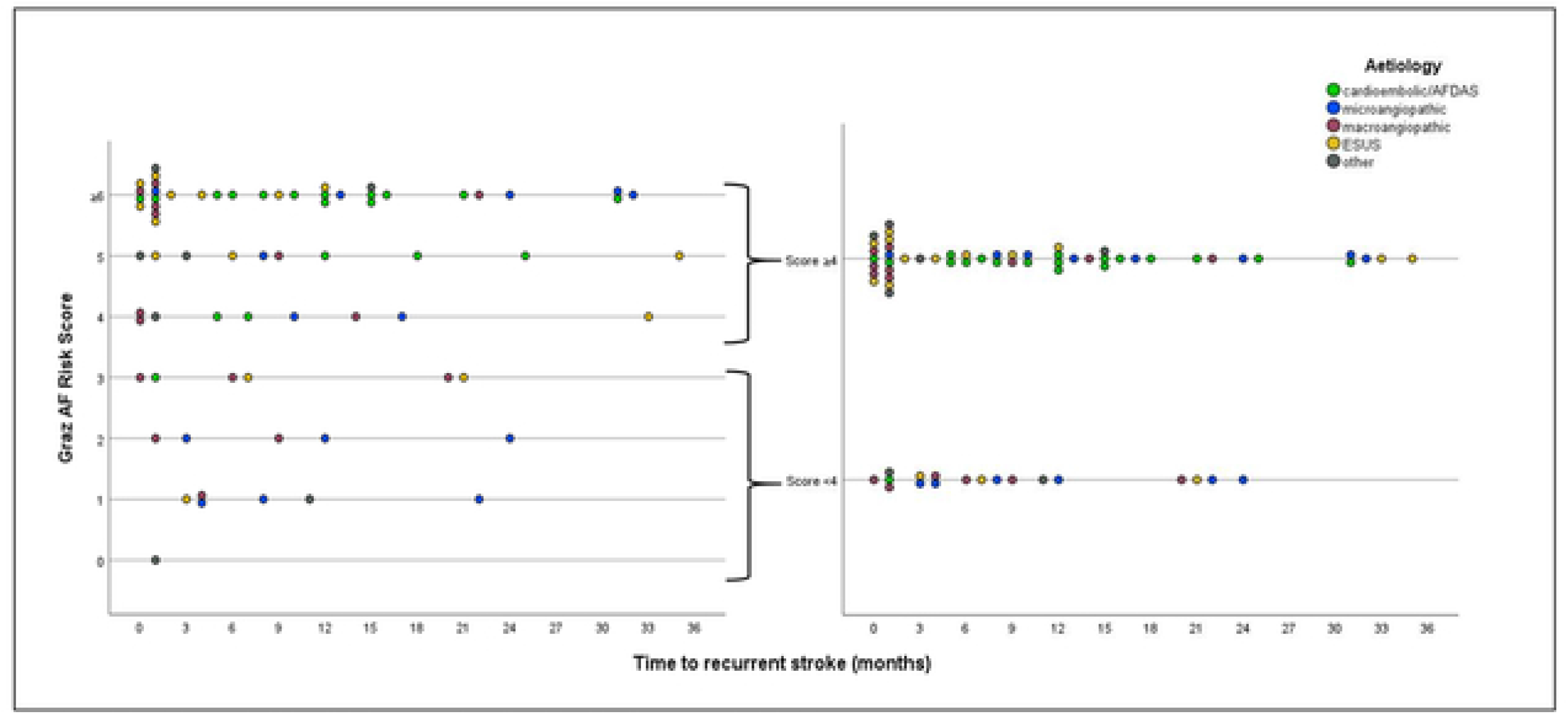
Recurrent stroke etiology according to Graz AF Risk Score and time to recurrence A, Distribution of recurrent stroke etiologies according to individual Graz AF Risk Scores and time from the index stroke to recurrent ischemic stroke. B, Distribution of recurrent stroke etiologies according to predefined Graz AF Risk Score category (<4 vs ≥4). Recurrent stroke etiology was classified as cardioembolic/AFDAS, macroangiopathic, microangiopathic, ESUS, or other. Abbreviations: AFDAS, atrial fibrillation detected after stroke; ESUS, embolic stroke of undetermined source.

**Table 3.** Clinical outcomes and recurrent stroke etiologies according to Graz AF Risk Score group.

| Outcome Variable | Score $\geq 4$<br>(n=396) | Score $< 4$<br>(n=388) | Absolute<br>risk<br>difference | HR<br>(95% CI) | p* |
| --- | --- | --- | --- | --- | --- |
| AFDAS (n, %) | 151 (38.1) | 15 (3.9) | 34.2 | 12.1<br>(7.1–20.6) | <.001 |
| Recurrent stroke (n, %) | 52 (13.1) | 18 (4.6) | 8.5 | 2.9<br>(1.7–5.0) | <.001 |
| Recurrent AF-related stroke<br>(n, %) | 14 (26.9) | 1 (5.6) | 21.3 | 14.3<br>(1.9–108.9) | <.001 |
| Recurrent non-AF related<br>cardioembolism (n, %) | 4 (7.7) | 0 (0.0) | 7.7 | - | .124 |
| Recurrent macroangiopathic<br>stroke (n, %) | 9 (17.3) | 6 (33.3) | -16.0 | 1.5<br>(0.5–4.3) | .458 |
| Recurrent microangiopathic<br>stroke (n, %) | 8 (15.4) | 6 (33.3) | -17.9 | 1.4<br>(0.5–3.9) | .616 |
| Recurrent ESUS (n, %) | 12 (23.1) | 3 (16.7) | 6.4 | 4.0<br>(1.1–14.0) | .021 |
| Recurrent stroke of other<br>etiology (n, %) | 5 (9.6) | 2 (11.1) | -1.5 | 2.5<br>(0.5–12.9) | .266 |
| Myocardial infarction (n, %) | 10 (2.5) | 3 (0.8) | 1.7 | 3.2<br>(0.9–11.5) | .055 |
| GI bleeding (n, %) | 8 (2.0) | 2 (0.5) | 1.5 | 3.8<br>(0.8–17.9) | .06 |
| All cause mortality (n, %) | 28 (7.1) | 3 (0.8) | 4.8 | 7.1<br>(2.1–23.9) | <.001 |

AFDAS, atrial fibrillation detected after stroke; ILR, implantable loop recorder; AF, atrial fibrillation; GI, gastrointestinal; HR, hazard ratio; CI, confidence interval; ESUS, embolic stroke of undetermined source
\*p values were calculated using Pearson's chi-square test or Fisher's exact test, as appropriate, for comparisons between patients with a Graz AF Risk Score $\geq 4$ and $< 4$

## Discussion

In this population-based study of consecutive patients with ESUS, implementation of the Graz AF Risk Score within a statewide risk-adapted diagnostic pathway enabled effective stratification of patients according to their risk of AFDAS. While AFDAS detection was low in patients with risk scores <4 points (3.9%), high-risk patients had a roughly ten-fold higher rate of AFDAS during the follow-up period (38.1%). Beyond AFDAS prediction, the score also identified patients at increased risk of recurrent ischemic stroke and differentiated distinct patterns of recurrent stroke mechanisms, supporting its value for risk stratification after ESUS.

Several prediction models have been proposed to guide AF screening in ESUS patients, but reported discrimination has been modest and derived from selected and small cohorts. ^25,27,33,34^ In contrast, the present study evaluated the Graz AF Risk Score within routine clinical practice across an entire regional stroke network. The observed AUC of 0.79 and the pronounced gradient in AFDAS detection across risk strata suggest that the score retains substantial discriminatory ability under real-world conditions.

The overall AFDAS detection rate of 21.2% was at the upper range of previously reported rates of ESUS cohorts, likely reflecting systematic rhythm surveillance and targeted allocation of intensive monitoring to patients with a higher pretest probability of AF. ^4,6,35–37^ In this context, the low number needed to monitor (2.9) in the high-risk group highlights the potential efficiency of a risk-adapted strategy.

Conversely, patients with scores below the predefined threshold rarely developed AFDAS during long-term follow-up, resulting in a negative predictive value of 96%. This suggests that the score may help identify patients in whom intensive monitoring is unlikely to provide substantial diagnostic yield and in whom alternative stroke mechanisms warrant particular attention.

Beyond AF detection, higher Graz AF Risk Scores were independently associated with recurrent ischemic stroke. Importantly, this relationship was not limited to AF detection itself, as recurrent stroke mechanisms differed according to risk category. Cardioembolism was the most frequent cause of recurrent stroke in high-risk patients, whereas recurrent events in low-risk patients were predominantly attributable to large-artery or small-vessel disease. Of note, only one patient in the low-risk group was diagnosed with AFDAS as the etiology of recurrent ischemic stroke despite more than two years of median follow-up.

These findings extend recent work emphasizing the uncertain relationship between AF detection and clinical outcomes by demonstrating clinically meaningful differences in recurrent stroke risk and stroke mechanisms across Graz AF Risk Score categories. ^12^ This distinction may have implications for future precision-medicine approaches in ESUS. Previous trials of empiric anticoagulation in ESUS populations failed to demonstrate benefit, likely reflecting the biological heterogeneity of the syndrome. ^7–10^ Our findings support the concept that markers of atrial disease may help identify patients in whom occult AF or atrial cardiopathy plays a dominant role, while simultaneously identifying a subgroup in whom alternative mechanisms deserve greater diagnostic focus.

## Limitations

Several limitations should be acknowledged. First, intensified cardiac rhythm monitoring was preferentially allocated to patients with higher Graz AF Risk Scores, resulting in substantially greater ILR implantation rates in the high-risk group. Consequently, part of the observed difference in AFDAS detection may reflect increased surveillance intensity. However, this pattern represents the intended implementation of the risk-adapted diagnostic pathway, and adjustment for ILR implantation did not materially alter the association between the score and AFDAS detection. Moreover, in the no-ILR group patients with scores ≥4 points had a 7.6-fold higher rate of AFDAS during the follow-up period compared to low-risk patients (25.1 vs 3.3%), but residual detection bias related to intensive rhythm monitoring cannot be excluded. Second, ILR implantation was systematically documented, whereas detailed information on the type and extent of non-ILR cardiac rhythm monitoring performed in individual patients was not consistently available. This may have influenced the observed AF detection rates. Third, despite comprehensive linkage of regional hospital information systems and the Austrian electronic health record infrastructure (ELGA), asymptomatic AF episodes that neither triggered clinical attention nor were detected during monitoring may have remained unrecognized. Finally, although the population-based design across an entire regional stroke network supports generalizability to routine stroke care, external validation in other healthcare systems and populations is warranted.

## Conclusion

The Graz AF Risk Score demonstrated robust performance for AFDAS detection in routine clinical practice. Beyond AFDAS prediction, the score identified clinically distinct risk groups with regard to recurrent stroke and recurrent stroke mechanisms. These findings support its use as a practical tool to facilitate risk-adapted cardiac rhythm monitoring in patients with ESUS.

## List of Abbreviations and Acronyms

AF: atrial fibrillation
AFDAS: atrial fibrillation detected after stroke
AUC: area under the curve
CI: confidence interval
ECG: electrocardiogram
EF: ejection fraction
ELGA: Austrian electronic health record system
ESUS: embolic stroke of undetermined source
GI: gastrointestinal
HR: hazard ratio
ILR: implantable loop recorder
IQR: interquartile range
NPV: negative predictive value
NT-proBNP: N-terminal pro–B-type natriuretic peptide
PPV: positive predictive value
ROC: receiver operating characteristic
SPB: supraventricular premature beats
SPSS: Statistical Package for the Social Sciences
STROBE: Strengthening the Reporting of Observational Studies in Epidemiology

## Authors’ contributions

JE and AK contributed equally to this work and share first authorship. JE and AK coordinated data collection, performed the statistical analyses, interpreted the data, prepared the figures and tables, and drafted the manuscript. TR contributed to data collection. SPKV performed data extraction from the electronic medical records.

TG, MK, MH, JJ, HK, SL, MM, EHB, DS, CE, MH, NB, IH, and SFH contributed to study implementation and patient recruitment at the participating centers and critically revised the manuscript for important intellectual content.

TG and MK supervised the implementation of the diagnostic pathway across the participating stroke centers. MH, JJ, HK, SL were responsible for implementation of the study pathway at the collaborating regional stroke centers. MM, EHB, DS provided cardiology expertise and contributed to study implementation. CE provided overall institutional oversight and critical scientific input.

MK conceived and supervised the study, interpreted the data, critically revised the manuscript, and had overall responsibility for the project.

All authors read and approved the final version of the manuscript.

## Data availability

Data from this study are available from the corresponding author upon reasonable request.

## Acknowledgements

OpenAI generative AI tools were used to create selected illustrative icons in Figure 1 and Supplemental Figure 1 and the artwork for the Graphical Abstract. All AI-assisted content was reviewed and approved by the authors.

## Supplemental Material

Supplemental Tables 1–7

Supplemental Figures 1–3

